# Gut metabolites predict *Clostridioides difficile* recurrence

**DOI:** 10.1101/2021.11.24.21266826

**Authors:** Jennifer J. Dawkins, Jessica R. Allegretti, Travis E. Gibson, Emma McClure, Mary Delaney, Lynn Bry, Georg K. Gerber

## Abstract

**Background:** *Clostridioides difficile* infection (CDI) is the most common hospital acquired infection in the U.S., with recurrence rates >15%. Although primary CDI has been extensively linked to gut microbial dysbiosis, less is known about the factors that promote or mitigate recurrence. Moreover, previous studies have not shown that microbial abundances in the gut measured by 16S rRNA amplicon sequencing alone can accurately predict CDI recurrence.

**Results:** We conducted a prospective, longitudinal study of 53 non-immunocompromised participants with primary CDI. Stool sample collection began pre-CDI antibiotic treatment at the time of diagnosis, and continued up to eight weeks post-antibiotic treatment, with weekly or twice weekly collections. Samples were analyzed using: (1) 16S rRNA amplicon sequencing, (2) liquid chromatography/mass-spectrometry metabolomics measuring 1387 annotated metabolites, and (3) short-chain fatty acid profiling. The amplicon sequencing data showed significantly delayed recovery of microbial diversity in recurrent participants, and depletion of key anaerobic taxa at multiple time-points, including *Clostridium* cluster XIVa and IV taxa. The metabolomic data also showed delayed recovery in recurrent participants, and moreover mapped to pathways suggesting distinct functional abnormalities in the microbiome or host, such as decreased microbial deconjugation activity, lowered levels of endocannabinoids, and elevated markers of host cell damage. Further, using predictive statistical/machine learning models, we demonstrated that the metabolomic data, but not the other data sources, can accurately predict future recurrence at one week (AUC 0.77 [0.71, 0.86; 95% interval]) and two weeks (AUC 0.77 [0.69, 0.85; 95% interval]) post-treatment for primary CDI.

**Conclusions:** The prospective, longitudinal and multi-omic nature of our CDI recurrence study allowed us to uncover previously unrecognized dynamics in the microbiome and host presaging recurrence, and, in particular, to elucidate changes in the understudied gut metabolome. Moreover, we demonstrated that a small set of metabolites can accurately predict future recurrence. Our findings have implications for development of diagnostic tests and treatments that could ultimately short-circuit the cycle of CDI recurrence, by providing candidate metabolic biomarkers for diagnostics development, as well as offering insights into the complex microbial and metabolic alterations that are protective or permissive for recurrence.

## Introduction

*Clostridioides difficile* infection (CDI) is the most common cause of health-care associated infection in the U.S., with symptoms ranging from diarrhea to life-threating fulminant colitis [1]. Annually in the U.S., there are >460K CDI cases and >30K deaths, with costs to the health care system estimated at >$4.8 billion [2]. CDI recurrence after initial infection is common, with an estimated overall 15.5% rate of first recurrence, and escalating recurrence risk with each subsequent episode [2], [3]. *Clostridioides difficile* is a Gram positive, anaerobic spore-forming bacteria that can colonize the gut asymptomatically, with estimates of asymptomatic colonization up to 17% of healthy adults in the community and 50% of hospital patients [1], [4]. Toxigenic strains of *C. difficile* can release endotoxins that bind to intestinal epithelial cells to cause cell death and severe inflammation [4], [5]. However, even toxigenic strains have been found to colonize asymptomatically, and dysbiosis of the microbiome is critical for CDI to occur [4]. Indeed, antibiotic exposure, particularly with drugs that deplete gut anaerobes, is a major risk factor for development of CDI [6], [7].

The mechanisms through which gut microbial dysbiosis drives CDI remain incompletely understood, but there is mounting evidence that the gut metabolome plays an important role. *C. difficile* is capable of metabolizing a variety of carbon sources, including proline, glycine, and branched-chain amino acids via Stickland fermentation [8]. Murine studies have shown that CDI decreases amino acid Stickland substrates and increases Stickland products such as 5-aminovalerate, indicating a utilization of Stickland substrates by *C. difficile* [9], [10]. In recent work in gnotobiotic mice, the commensal bacteria *Paraclostridium bifermentans*, which preferentially uses Stickland fermentation for energy and depletes Stickland substrates in the gut, provides strong protection against CDI infection [11]. Certain cholate-derived primary bile acids, which are depleted in a healthy gut microbiome due to microbial metabolism, have been shown to be co-germinants for *C. difficile in vitro*. However, the role of these metabolites *in vivo* is less clear, and recent studies have shown that the mechanism by which microbes such as *Clostridium scindens* provide protection *in vivo* may be due to their utilization of *C. difficile’s* preferred carbon sources, rather than through primary bile acid depletion [11]–[13]. Short chain fatty acids (SCFAs) have also been associated with CDI, although their role is less clear. Acetate and butyrate, gut microbial products of dietary fiber fermentation, have been associated with general gut health in some studies; butyrate, in particular, is a primary energy source for colonocytes and thus may help maintain intestinal barrier integrity [14]. However, *Clostridium sardiniense*, which significantly increases butyrate in the gut, was not protective against CDI in gnotobiotic animal studies, and in fact worsened infection [11]. Taken together, evidence drawn from *in vitro* or murine studies suggests that CDI may be driven by a multifactorial gut metabolic dysbiosis, which includes alterations in carbon sources.

Despite compelling evidence for the importance of gut metabolomic dysbiosis in CDI, to our knowledge, there have only been three studies that analyzed metabolic factors of CDI in reasonably sized (>20 subjects) human cohorts. Allegretti et al. performed a cross-sectional comparison of bile acid profiles of participants with first-time CDI (n=20), recurrent CDI (n=19), and no CDI (n=21), and found higher primary bile acids and lower secondary bile acids in those with CDI versus those without CDI [15]. Robinson et al performed a cross-sectional analysis of untargeted metabolomes of participants (n=186) with CDI versus with non-CDI diarrhea, and found higher Stickland fermentation products and lower fructose in CDI participants [16]. Bushman et al. compared the metabolomes of children with IBD (n=27), children with IBD and CDI (n=23), and healthy controls (n=38) at CDI diagnosis, 4 weeks, and 8 weeks later, and found higher primary bile acids, sphingomyelins, and intracellular fatty acids in CDI+IBD and IBD children [17].

CDI recurrence has also been relatively understudied, and it remains unclear whether the factors described above for primary CDI play similar roles in recurrent disease. Pakpour et al. assessed whether the composition of the gut microbiome could predict recurrence, but found only a weak relationship (area under the receiver-operator curve [AUC] of 0.61) [18]. Four other studies have investigated predicting recurrence solely using electronic health record (EHR) data, and have achieved AUCs ranging from 0.67 to 0.82 [19]–[22]. Three of these studies found proton-pump inhibitor use to be predictive of recurrence, and two of the studies found higher age to be predictive of recurrence; however, there were no other predictive features common among the studies. Moreover, two of these studies on independent cohorts was attempted, and found poor predictive accuracy [23].

To address the gaps in prior studies, including cross-sectional analyses, lack of metabolomic data, and potentially confounding comorbidities or antibiotic use, we conducted a prospective study in which we collected longitudinal stool samples from participants without IBD or immunocompromise, from the time of CDI diagnosis prior to initiation of antibiotic treatment, and up to eight weeks post-treatment (or until the time of recurrence). Samples were interrogated via broad LC/MS metabolomic profiling, 16S rRNA amplicon sequencing, and targeted short-chain-fatty-acid (SCFA) analysis. We used univariate and multivariate statistical techniques to investigate how microbial composition and metabolomes of recurrers vs. non-recurrers changed and diverged over time. Further, we used cross-validated machine learning/statistical methods to quantify the capability of the data sources to predict future recurrences.

## Results

### Longitudinal study of recurrent CDI measuring gut microbiome and metabolome

We conducted a prospective, longitudinal study of 53 participants on inpatients at Brigham and Woman’s Hospital’s, as well as two surrounding community hospitals (Figure 1). Non-immunocompromised participants experiencing uncomplicated CDI were followed for up to eight weeks after completion of their CDI antibiotic treatment or until recurrence. During this time, 19 participants were diagnosed with recurrent CDI, with all recurrences within the first three weeks post-CDI antibiotic treatment. Diagnosis used a two-step testing algorithm, glutamate dehydrogenase (GDH) reflexed to enzyme immunoassay (EIA) toxin testing. Table 1 provides demographic information on participants’ sex, race, smoking status, age, and BMI; there were no significant differences in any of these variables with respect to CDI recurrence status.

**Fig 1.**
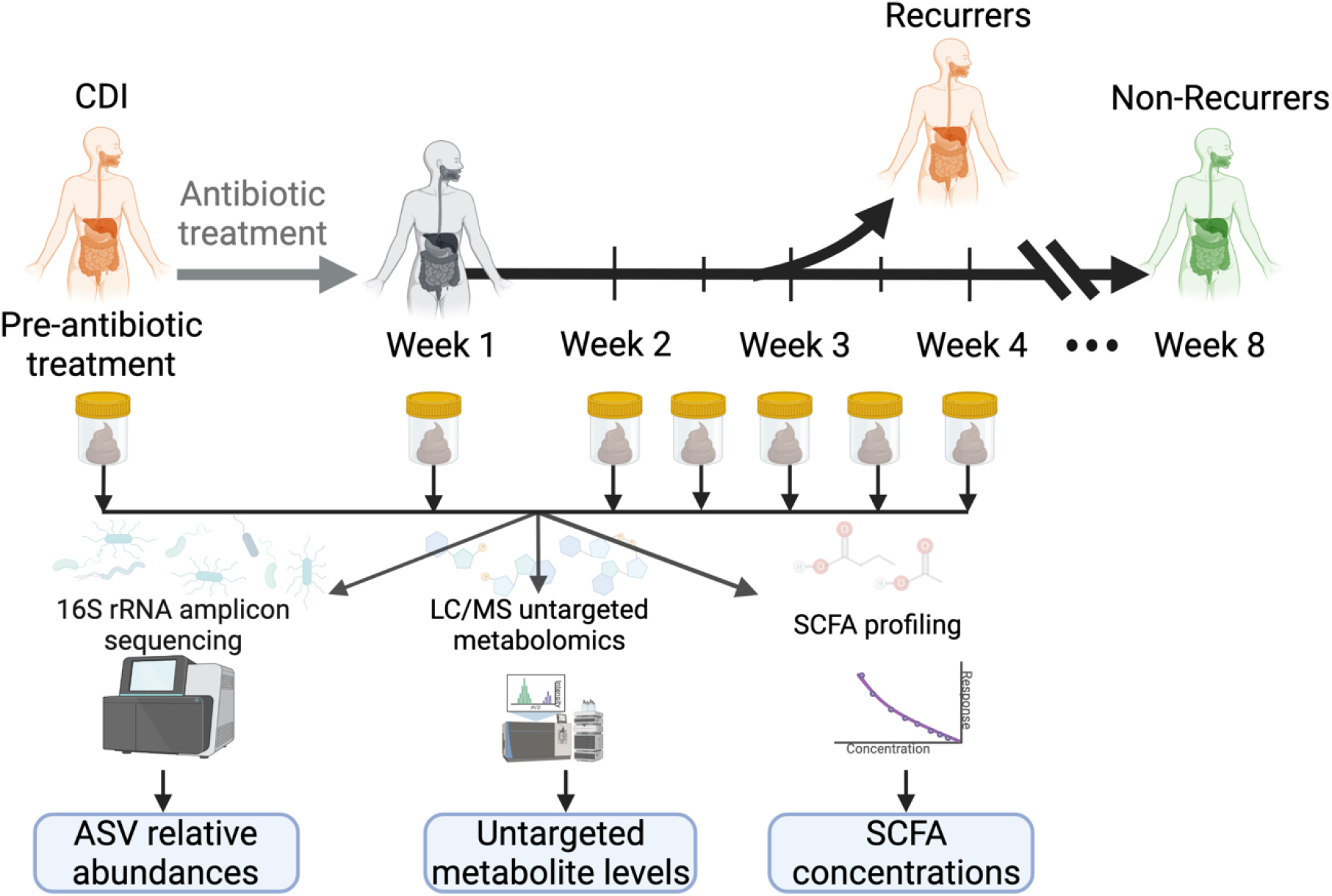
Prospective study of *Clostridioides difficile* infection (CDI) measured gut microbiome composition and metabolomes for developing predictors of recurrence. Fifty-three participants with first-time CDI were followed for up to eight weeks after initial CDI antibiotic treatment. Fecal samples were collected prior to CDI antibiotic treatment, one week post-treatment, and then weekly or bi-weekly until recurrence or end of the study period. Microbial composition within fecal samples was analyzed with 16s rRNA gene amplicon sequencing. Metabolites in fecal samples were measured with liquid chromatography/mass spectrometry (LC/MS) broad metabolomics and targeted short chain fatty acid profiling.

**Table 1.** Participant demographics. Statistical testing was performed using Fisher’s exact test for binary variables, the chi-squared test for categorical variables, and the Wilcoxon rank-sum test for continuous variables.

|  |  | Non-Recurrers | Recurrers | p-value |
| --- | --- | --- | --- | --- |
| Sex | Male | 13 | 5 | 0.226 |
|  | Female | 21 | 14 |  |
| Race | Black | 7 | 1 | 0.824 |
|  | White | 25 | 15 |  |
|  | Hispanic | 2 | 3 |  |
| Smoking status | Never | 31 | 13 | 0.612 |
|  | Former | 20 | 5 |  |
|  | Current | 2 | 1 |  |
| Age | Mean | 57.6 ± 15.6 |  | 0.636 |
|  | Median | 57 |  |  |
|  | Range | 22-93 |  |  |
| BMI | Mean | 28.9 ± 8.49 |  | 0.875 |
|  | Median | 26.3 |  |  |
|  | Range | 19.4-66.7 |  |  |

Fecal samples were collected at the time of diagnosis (if available), one week after antibiotic treatment, and every week or half week for up to eight weeks, or until recurrence (Table 2). Because all participants who recurred did so within the first three weeks after initial treatment, we focused our subsequent analyses primarily on data-points prior to week three. This time window provides a sufficient number of recurrent samples for statistical testing and also, in the context of developing diagnostic testing in the future, represents a relevant time period for clinically actionable decision-making. Each sample was analyzed with: (1) 16S rRNA amplicon sequencing, (2) liquid-chromatography/mass-spectrometry (LC/MS) untargeted metabolomics, and (3) targeted short chain fatty acid (SCFA) analyses. For amplicon analyses, this yielded 4,605,740 total sequencing reads (average of ∼10K/reads per sample) and subsequent bioinformatic processing with dada2 produced 2509 unique amplicon sequence variants (ASVs). The LC/MS untargeted metabolomics platform quantified 1387 unique and annotated metabolites. SCFA analyses quantified nine metabolites: acetate, propionate, isobutyrate, butyrate, isovalerate/2-methylbutyrate (indistinguishable by the platform used), valerate, isocaproate, caproate, and heptanoate. However, heptanoate and caproate were only present in one or two samples, respectively, and were thus removed from subsequent analyses.

**Table 2.** Number of samples available from non-recurrers and recurrers at each time-point.

| Week | Non-recrurers | Recurrers |
| --- | --- | --- |
| Pre-CDI antibiotics | 18 | 8 |
| 1 | 34 | 14 |
| 1.5 | 10 | 3 |
| 2 | 34 | 6 |
| 2.5 | 10 | 2 |
| 3 | 33 | 3 |
| 3.5 | 4 | 0 |
| 4 | 34 | 0 |

### Participants who recurred exhibited slower recovery of gut microbiome diversity and composition post-CDI antibiotic treatment

We first investigated differences in overall ecological diversity of the gut microbiome using alpha and beta diversity measures. As expected, alpha diversity [24] significantly decreased pre- to one week post-CDI antibiotic treatment within both recurrent (*p*=0.04) and non-recurrent (*p*=2×10^−4^) groups (Figure 2; Table S1), consistent with depletion of gut microbes during antibiotic treatment for CDI. Interestingly, over the following week, alpha diversity recovered significantly only within the non-recurrent group (*p*=3×10^−5^). Comparing alpha diversity between the recurrent and non-recurrent groups at each time-point, we found only a significant difference at week two post-CDI antibiotic treatment, with higher diversity in the non-recurrent group (*p*=2×10^−3^). We evaluated beta diversity using the Bray-Curtis dissimilarity measure and found a similar pattern (Figure S1). Significant changes in beta diversity from pre- to one week post-CDI antibiotic treatment were seen in both recurrers (*p*=5×10^−3^) and non-recurrers (*p*=10^−3^), but only non-recurrers had significant changes in beta diversity after CDI antibiotic treatment from week one to week two (*p*=10^−3^) (Table S1). Taken together, these results suggest that recurrent and non-recurrent participants both had expected declines in gut microbiome ecological diversity with antibiotic treatment for CDI, but recurrent subjects exhibited delayed recovery of microbial diversity.

**Fig 2.**
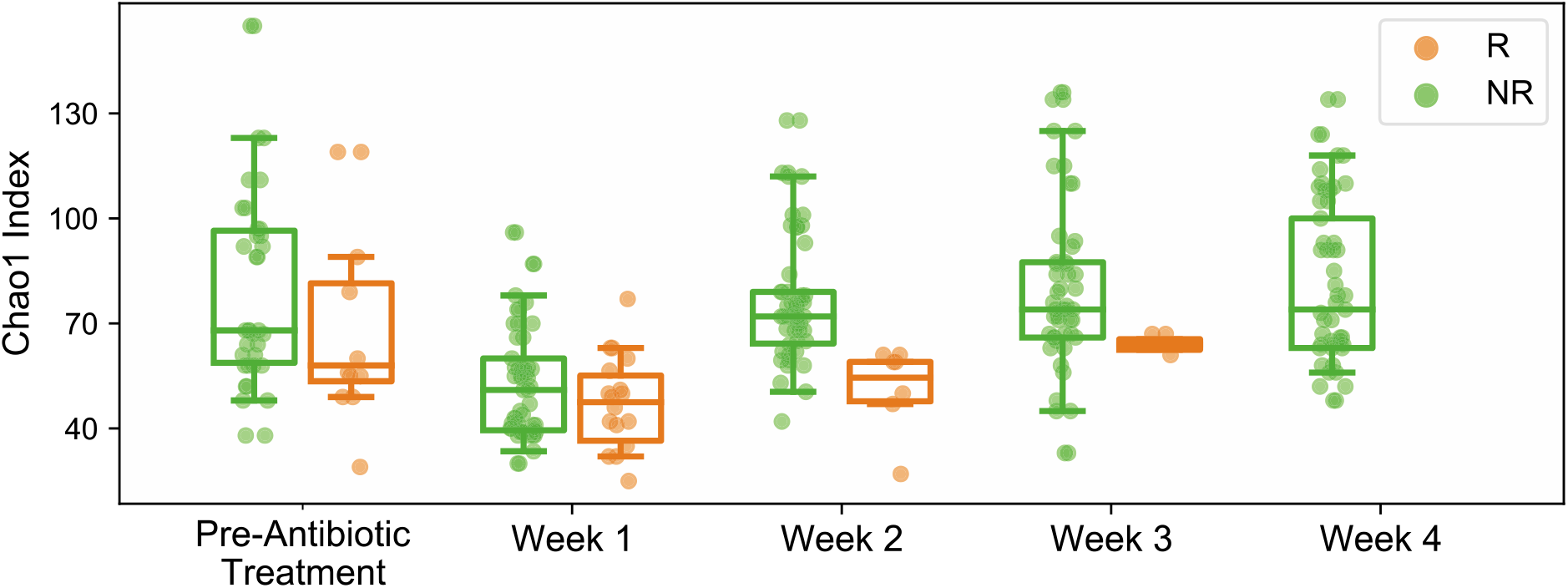
Ecological diversity of gut microbiomes of CDI recurrent participants recovered significantly more slowly than for non-recurrent participants. Alpha diversity (Chao index), a measure of species richness, significantly decreased pre- to one week post-CDI antibiotic treatment within both recurrent and non-recurrent groups. From one week to two weeks post-CDI treatment, alpha diversity recovered significantly only within the non-recurrent group. Alpha diversity only differed significantly between the recurrent and non-recurrent groups at two weeks post-CDI antibiotic treatment, with higher diversity in the non-recurrent group. R=Recurrers, NR=Non-recurrers.

We next examined differences in microbiome composition at the level of ASVs. After filtering low abundance/rare taxa, we obtained 237 ASVs, which we used for subsequent DESeq2 fold-change analyses. Analyzing changes in composition over time, we found that in non-recurrers, 12 ASVs significantly differed in abundance between week one to week two. Among these 12 ASVs, 10 exhibited significant increases (Table S2). This set of ASVs was significantly enriched for ASVs in the Lachnospiraceae family (*Fusicatenibacter saccharivorans* [ASV 83], *Roseburia inulinivorans* [ASV 205], a species within the *Ruminococcus* genera [ASV 212], and three *Clostridium* cluster XIa taxa [ASVs 90, 98, and 214]) (FDR=0.04), which are generally strict anaerobes with specialized niches and associated with normal microbiome function. In recurrers, 8 ASVs were significantly different in abundance from week one to week two; 5 of these ASVs significantly decreased, including *Akkermansia muciniphila* (ASV 10), which significantly increased in the non-recurrers over the same time-period. Comparison of microbial composition between recurrers and non-recurrers revealed that non-recurrers had significantly higher abundances of 10 ASVs pre-CDI treatment, 8 ASVs at week one post-CDI treatment, and 29 ASVs at week two (Figure 3A; Table S2). The set of ASVs at increased abundance at week two was significantly enriched for taxa in the Ruminococcaceae (FDR=0.03) family, and border-line significant for enrichment of Lachnospiraceae (FDR=0.06), and Bacteroidaceae (FDR=0.06). Many taxa in these families have been associated with normal microbiome function, including *Faecalibacterium* (ASV 4), *Subdoligranulum* (ASV 1), *Anaerotruncus* (ASV 110), *Oscillibacter* (ASV 173), and *Clostridium* cluster IV (ASVs 59, 66) taxa in the Ruminococcaceae family; *Blautia* (ASVs 102, 76, 77, 82), *Roseburia* (ASVs 205), *Clostridium* cluster XIVa taxa (ASVs 214, 99), *Anaerostipes* (ASVs 72), and *Fusicatenibacter* (ASV 83) taxa within the Lachnospiraceae family; and *Bacteroides* (ASVs 33,34, 29, 26) within the Bacteroidaceae family. Interestingly, one of the *Clostridium* cluster XIVa taxa at higher abundance in non-recurrers (significant at week two and with a trend toward higher abundance at other time-points) was *Clostridium scindens* (ASV 99), which has been shown to provide host resistance to *C. difficile* [12], [13]. A number of the other genera found to be at higher abundance in non-recurrers have been previously linked to protection against CDI in human studies, including *Bacteroides* (ASVs 26, 29, 33) and *Veillonella* (ASV 154) [18], [25]. Taken together, these results suggest a picture of broader depletions of the normal microbiome in recurrers, evident even pre-CDI antibiotic treatment, but with increasingly more pronounced differences over time, consistent with slower recovery of recurrers’ microbiomes.

**Fig 3.**
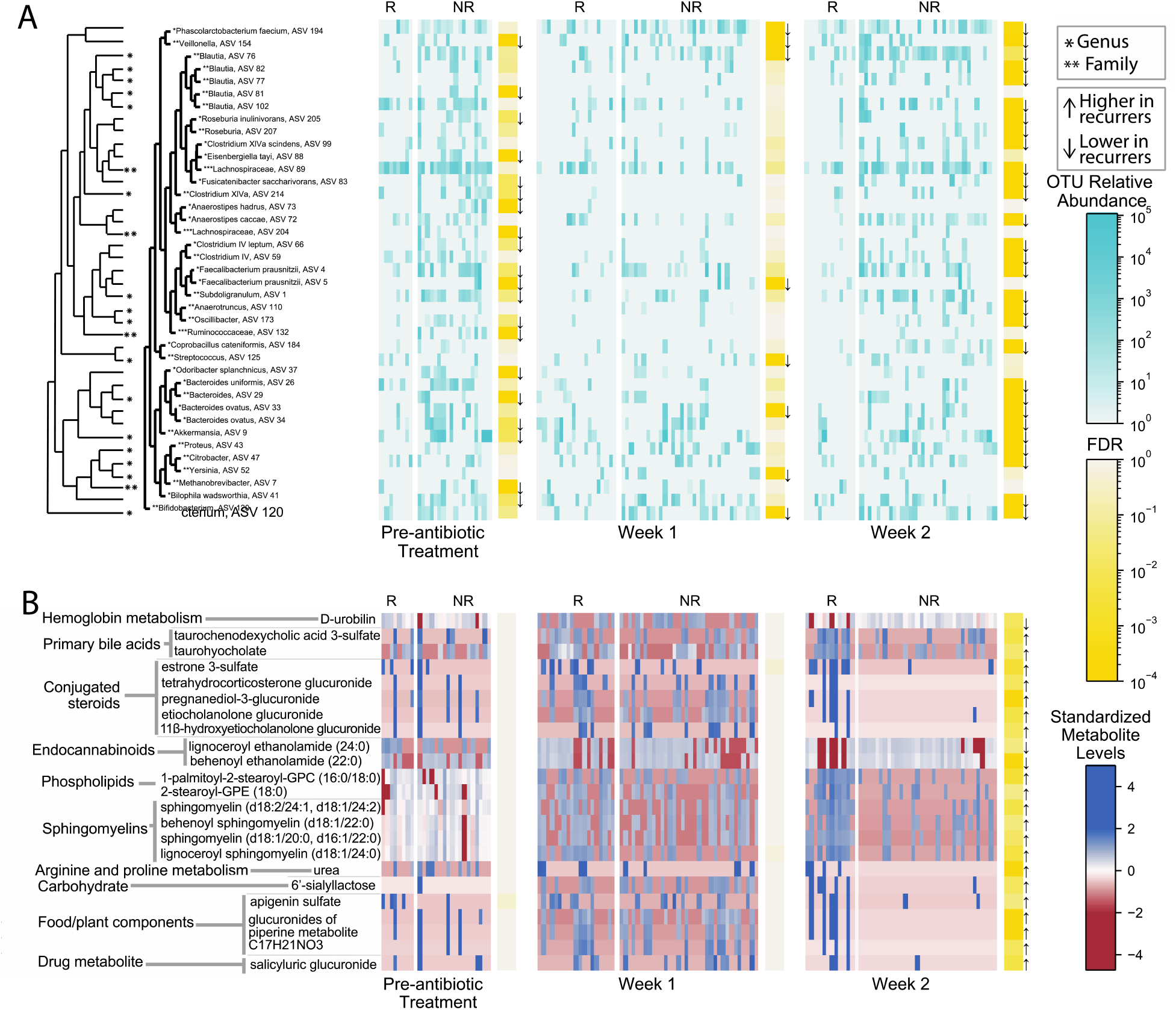
Gut microbiome taxa and metabolite levels differed significantly between CDI recurrent and non-recurrent participants. **(A)** Univariate analyses of 16S rRNA gene amplicon sequencing data found 39 out of 237 amplicon sequencing variants (ASVs) (post-filtering to remove rare or low-variance taxa), were significantly differentially abundant between recurrers versus non-recurrers. **(B)** Univariate analyses of LC/MS untargeted metabolomics found 23 out of 1387 metabolites (post-filtering to remove rare or low-variance metabolites), were significantly differentially abundant between recurrers versus non-recurrers. Metabolite levels shown are log-transformed and standardized. R=Recurrers, NR=Non-recurrers. Arrows denote the direction of the statistically significant effect. Participants (columns) were ordered in the figure via hierarchical clustering.

### Participants who recurred exhibited an altered gut metabolome indicative of reduced gut microbiome function, and host inflammation and reduced immune modulatory capabilities

We first performed ordination analyses to evaluate overall changes and differences in broad gut metabolomic profiles between recurrers and non-recurrers (Figure S2). Paralleling our findings on changes in microbial diversity, ordination analyses on metabolomic data (760 metabolites after filtering) showed that the metabolomes of non-recurrers changed significantly from pre-CDI treatment to week one post-CDI treatment (*p*=10^−3^), and from week one to week two post-CDI treatment (*p*=10^−3^), but the metabolomes of recurrers only changed significantly from pre-CDI treatment to week one post-CDI treatment (*p*=10^−3^) (Table S1). Comparing recurrers to non-recurrers at each time-point, differences were only significant at week two (*p*=0.001), which recapitulated our findings in microbiome alpha diversity (Table S1). Taken together, we saw parallel patterns for overall gut metabolomic profiles and microbial diversity, with recurrers and non-recurrers initially exhibiting similar gut metabolomes that only significantly diverge by week two post-CDI antibiotic treatment, due to a slower recovery in the recurrent group.

To determine which gut metabolites contributed to these overall patterns of metabolome recovery or non-recovery, we performed univariate analyses, both across time and between recurrers and non-recurrers on broad metabolomic data (Figure 3B, Table S2) and targeted SCFA data (Figure S3, Table S2). Changes in metabolites over time were significant only for non-recurrers from week one to week two, with 121 metabolites significantly changing over the week. These metabolites were significantly enriched for secondary bile acids (FDR=0.04), primary bile acids (FDR=0.04), and hydroxy acyl carnitines (FDR=0.04), and borderline significant for glucuronidated corticosteroids (FDR=0.07) and metabolites involved in hemoglobin and porphyrin metabolism (FDR=0.07). Changes in SCFAs over time (Table S2) were similarly only significant for non-recurrers, from week one to week two, with six SCFAs significantly higher: acetate (FDR=9×10^−5^), isovalerate/2-methylbutyrate (FDR=4×10^−4^), butyrate (FDR=10^−3^), valerate (FDR=0.03), and isobutyrate (FDR=0.03). Comparison between levels of gut metabolites in recurrers versus non-recurrers showed increasing differences over the study. At pre-treatment and week one post-treatment, no metabolites were found to be significantly different between recurrers and non-recurrers. However, at week one post-CDI treatment, two metabolites were at higher abundances in recurrers with borderline significance: vanillylmandelate (FDR=0.08) and N-carbamoylaspartate (FDR=0.08). At week two, abundances of 23 metabolites differed significantly between recurrers and non-recurrers (Table S2), with 20 of these metabolites showing higher levels in recurrers. This pattern of increasing divergence over time between gut metabolomes of recurrers and non-recurrers parallels the pattern seen with microbiome composition, suggesting slower recovery of the gut metabolome in recurrers.

The specific changes or differences in metabolites observed can generally be organized into three categories indicative of: (1) host inflammation or intestinal damage, (2) lack of microbial deconjugation activity, (3) host alterations in immune and inflammatory capability. Vanillylmandelate (VMA), higher in recurrers at week one post-CDI treatment, is an end product of catecholamine metabolism and has been previously reported as a biomarker of inflammation [26]. By week two post-CDI treatment, biomarkers of cell death were significantly elevated in recurrers. The overall set of metabolites differentiating recurrers and non-recurrers at week two was significantly enriched for sphingomyelins (FDR=5×10^−4^), including lignoceroyl sphingomyelin d18:1/24:0, sphingomyelin d18:2/24:1, d18:1/24:2, sphingomyelin d18:1/20:0, d16:1/22:0, and behenoyl sphingomyelin d18:1/22:0. In additional to sphingomyelins, two phospholipids, palmitoyl-2-stearoyl-GPC 16:0/18:0 and 2-stearoyl-GPE 18:0, were also significantly higher in recurrers. Elevated sphingomyelin and phospholipid metabolites have previously been associated with active intestinal epithelial damage, such as in murine models of CDI and in humans with CDI or IBD [10], [17].

At week two, evidence of impaired microbial function in the gut was also present in recurrers’ metabolomes. Glucuronide and sulfate conjugates were significantly higher in recurrers, including five steroid conjugates (pregnanediol-3-glucorinide, estrone 3-sulfate, 11 beta-hydroxyetiocholanolone glucuronide, etiocholanolone glucuronide, and tetrahydrocorticosterone glucuronide) and four additional glucuronidated compounds (three glucuronides of piperine metabolite C17H21NO3 and salicyluric glucuronide). Gut microbes are critical for deconjugation activities [27], [28]; thus, elevated levels of conjugated metabolites in recurrers may indicate significantly blunted recovery of this normal microbiome function. The microbiome is also critical for transforming bile acids. Indeed, two bile acids, taurocholate [FDR=0.005] (a primary bile acid) and taurochenodeoxycholic acid 3-sulfate [FDR=0.02] (a primary bile acid conjugate), were significantly higher in recurrers, again suggesting delayed recovery of microbiome function. Interestingly, taurocholate and other cholate derivatives have been demonstrated to promote *C. difficile* germination *in vivo*, though their role in pathogenesis *in vitro* is less clear [1], [11], [13], [29], [30]. Bilirubin metabolism is another major function of the gut microbiota [31]. D-urobilin, the end product of bilirubin metabolism, was significantly lower in recurrers [FDR=0.01]. Higher levels of SCFAs indicate active microbiota metabolism in the gut [14]. Consistent with the picture of gut microbial dysbiosis seen with the other metabolites discussed above, levels of acetate (FDR=0.07) and isovalerate/2-methylbutyrate (FDR=0.7) were borderline significant for being lower in recurrers.

At week two, levels of metabolites involved in host immune or inflammatory modulation, predominately conjugated anti-inflammatory compounds and endocannabinoids, also differed significantly between recurrers and non-recurrers. The observed lower levels of conjugated corticosteroids in non-recurrers not only indicates greater microbial deconjugation activities, but may also indicate increased host anti-inflammatory activity: unconjugated corticosteroids, such as tetrahydrocorticosterone, are key anti-inflammatory compounds, and unconjugated sex steroids have also been shown to act as important modulators of inflammation in the gut [28], [32]. Other conjugated compounds found to be significantly higher in recurrers, specifically glucuronides of piperine, salicyluric glucuronide and apigenin sulfate, have also been shown to have unconjugated forms with anti-inflammatory effects [33]–[36]. Levels of the endocannabinoids behenoyl ethanolamide and lignoceroyl ethanolamide were also significantly lower in recurrers. Endocannabinoids have been shown to maintain gut homeostasis through modulating the immune system and gut motility; additionally, endocannabinoids have been found to increase in the presence of *Akkermansia muciniphila*, a taxa we found to be significantly more abundant in non-recurrers at week two [37]–[39]. Taken together, these results suggest a picture of reduced capability to modulate inflammation in recurrers.

### Predictive models of recurrence achieved highest accuracy using metabolomic data

To estimate how well our data can predict CDI recurrence in patients, we built supervised machine learning/statistical models and evaluated them using cross-validation. This approach fundamentally differs from the univariate statistical tests presented in the previous sections in two ways: (1) univariate approaches evaluate one variable at a time, and thus cannot find combined effects (e.g., increased risk if multiple metabolites are elevated), and (2) statistical testing approaches cannot provide an estimate of predictive accuracy, or how well the model might perform on unseen data. Both these capabilities are necessary for developing a clinically useful diagnostic, which is an important objective in the field. For prediction tasks, we evaluated three standard methods in the field: lasso-logistic regression (LR), random forests (RF), and lasso-Cox regression (CR). The first two methods predict binary outcomes (recurrence or non-recurrence), whereas CR predicts the time to recurrence. We evaluated these methods based on their ability to predict outcomes using a cross-validation methodology (training the models on subsets of the data and predicting on held-out data). For the two methods predicting binary outcomes, we used the area under the receiver operator curve (AUC) score as the evaluation metric, and for CR we used the concordance index (CI).

We applied LR, RF, and CR to participants’ pre-CDI treatment, week 1, and week 2 samples, using the following information: (1) clinical variables found to be associated with recurrence in prior studies (age, previous PPI use, treatment regiment, and diagnosis method), (2) ASVs from 16S rRNA amplicon sequencing, (3) LC/MS untargeted metabolomics, (4) SCFAs, and, (5) all sources of data (1-4) combined. Overall, we found that the LC/MS metabolomic data at weeks one and two had the highest predictive accuracy (Fig 4; Table S4). For predicting recurrence/non-recurrence, at week one, LR on metabolomic data achieved the highest AUC (0.77; [0.71, 0.86] 95% interval), and at week two, RF on metabolomic data achieved the highest AUC (0.77; [0.69, 0.85] 95% interval). None of the other data sources or time-points achieved AUC scores greater than 0.7, which is generally considered the threshold for an acceptable clinical test (with 0.8 to 0.9 considered excellent). Models predicting recurrence using all available data sources combined achieved essentially equivalent AUCs to models using only metabolomics data (Figure 4); moreover, these models only consistently selected metabolites as the significant features needed to make predictions (Table S5). Prediction of survival time using CR followed similar trends, as all models that achieved CIs > 0.7 selected only metabolites to make predictions. Both ASVs and SCFAs at pre-CDI treatment achieved median AUCs close to 0.7 (0.68 using LR for ASVs, and 0.68 using RF for SCFAs). However, the 95% cross-validated intervals for these AUCs were large, with their lower ranges extending toward values near 0.5 (random chance). Thus, these predictors lack robustness or generalizability. The lack of accurate pre-treatment predictors may have been limited by sample sizes in our study, as fewer samples were available pre-CDI treatment (N=26), compared to weeks one and two (N=48 and N=40, respectively).

**Fig 4.**
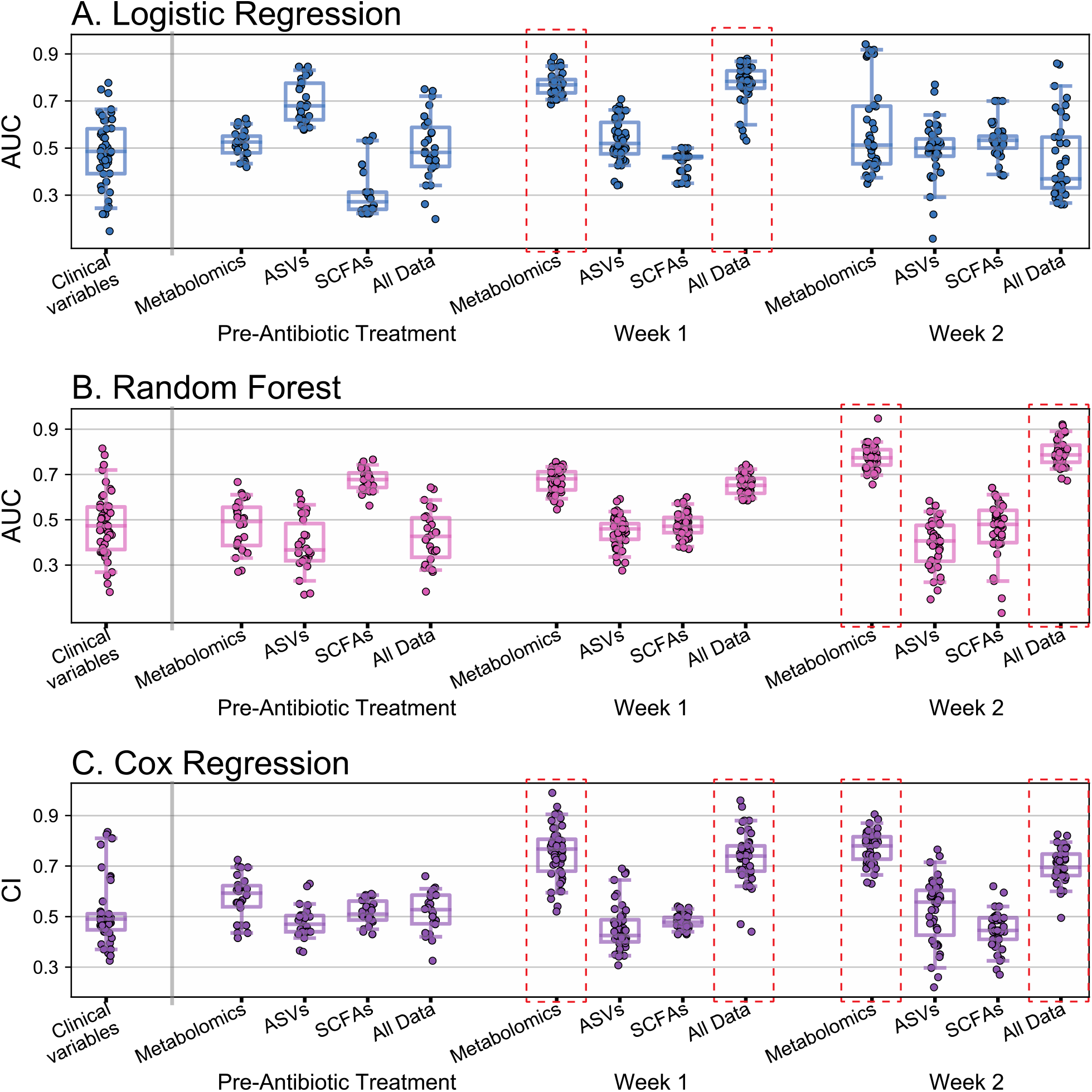
Predictive modeling of CDI recurrence achieved the highest accuracy using metabolomic data. The performance of predictive models was assessed using leave-one-out cross-validation (N=26 at pre-CDI treatment, N=48 at week one, and N=40 at week two). Data sources input to models were: (1) clinical variables associated with recurrence in prior studies (age, previous PPI use, antibiotic treatment regimen, and CDI diagnostic test used), (2) untargeted gut metabolomics, (3) amplicon sequencing variants (ASVs) of the gut microbiome, (4) gut short chain fatty acids (SCFAs), (5) data sources 1-4 combined. Performance of **(A)** logistic regression with lasso and **(B)** random forests, which predict binary labels (recurrence/no recurrence), were assessed with the area-under-the-curve (AUC)metric. **(C)** Cox regression, which predicts survival time, was assessed with the concordance index (CI). Models achieving median ≥0.70 AUC or CI scores (adequate performance) are denoted with red dashed rectangles. The “All Data” models with ≥0.70 AUC or CI were found to select only metabolomic features.

To determine which metabolites were predictive in models with median AUCs > 0.7, we assessed cross-validated odds ratios and Gini feature importance measures. At week one, LR, RF and CR all selected N-carbamoylaspartate and vanillylmandelate as the top predictors, both of which favored recurrence when at higher levels (Figure 5). Of note, these metabolites were also found in univariate analysis to be borderline-significantly increased in recurrers at week one. At week two, RF robustly identified lignoceroyl sphingomyelin as an important feature; this metabolite was also found to be significantly more abundant in recurrers in univariate analyses. RF also identified features with borderline significance that were found in the univariate analyses, including sphingomyelins, primary bile acids, and a phosphorylated lipid (Figure 5). The predictive models also identified features that were not detected in univariate analyses: 4-hydroxyhippurate and bilirubin in the week one LR model were identified as predictive of recurrence when at higher levels. 4-hydroxyhippurate is a product of microbial degradation of polyphenols found in fruits and other plant-based foods [40]. Bilirubin is the product of host heme catabolism and is further reduced to urobilinoids/urobilinogens by the gut microbiome, so its higher levels in recurrers’ gut metabolomes is consistent with subpar microbiome function [31]. Because the predictive methods employed make different underlying assumptions (e.g., logistic regression is a generalized linear model whereas random forests is a black-box nonlinear model), metabolites selected by multiple models are more likely to be robust [41]. Thus, the set of predictive metabolites identified by multiple methods (Figure 5) may serve as strong candidates for future trials to validate biomarkers for recurrence prediction in larger, independent cohorts.

**Figure 5.**
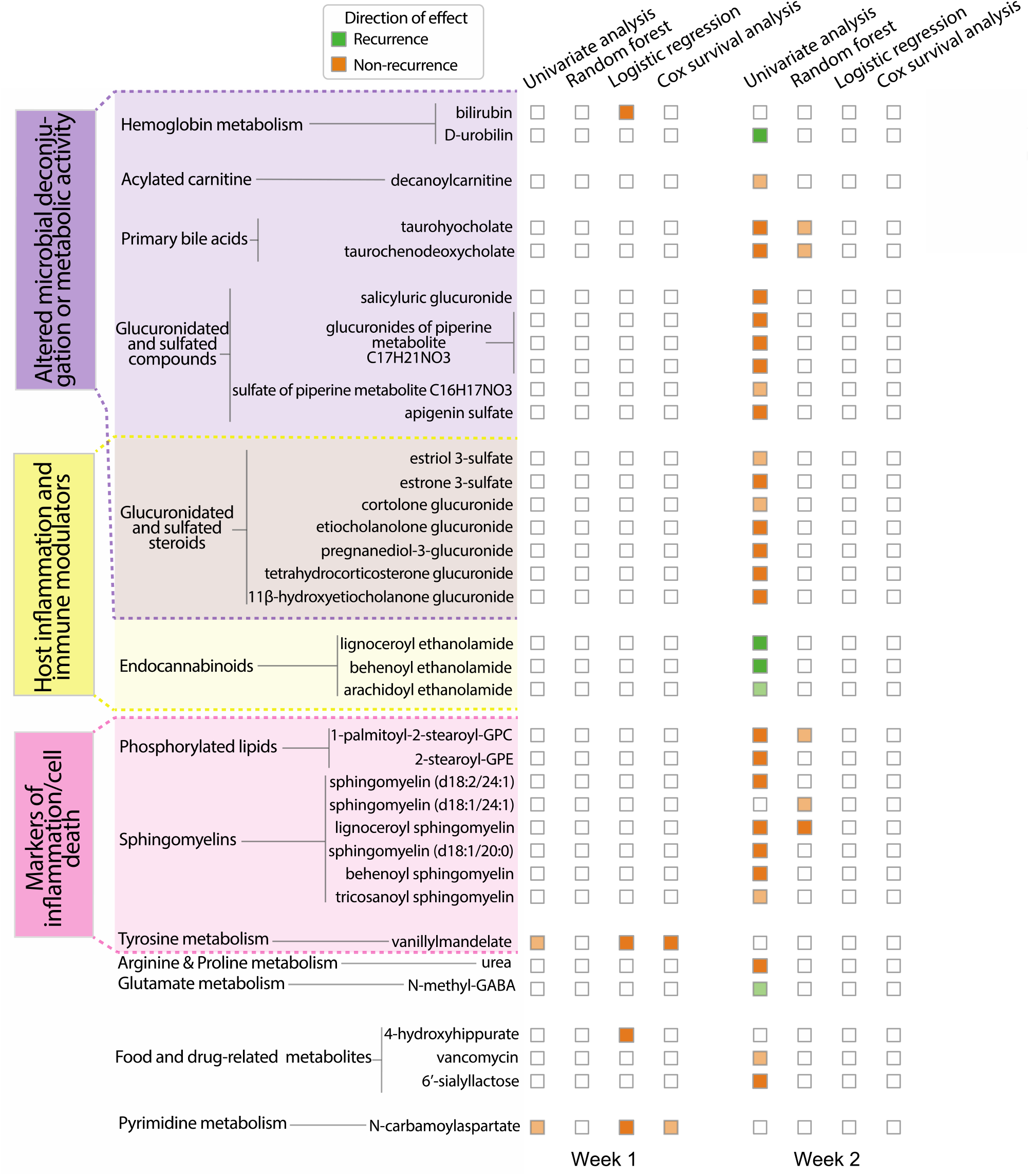
Integration of results of multiple analysis methods revealed that alterations in gut metabolites mapping to distinct host or microbiome-associated processes presaged CDI recurrence. Thirty-seven metabolites were significant in at least one analysis method for distinguishing CDI recurrent versus non-current status. These metabolites fell into one of three categories, reflecting altered host or microbiome activities. Dark orange or green colors indicate significance (FDR<0.05 in univariate analyses; 95% cross-validated log-odds/feature importance interval not containing 1.0 for predictive models). Light orange or green colors indicate borderline significance (0.05<FDR≤0.10 in univariate analyses; 75% log-odds/feature importance cross-validated interval not containing 1.0 for predictive models).

## Discussion

To our knowledge, our work represents the largest prospective study of CDI recurrence employing both microbiome sequencing and untargeted metabolomics analyses. Although prior studies have investigated some aspects of the relationship between the microbiome and CDI, our study design and analysis methods allowed us to probe further. The longitudinal nature of our study allowed us to investigate how rates of microbiome recovery relate to recurrence. Past studies [15]–[17] were cross-sectional or did not sample systematically in the period following antibiotic treatment for CDI, which is when we observed the most marked divergence between recurrers and non-recurrers. Importantly, the prospective nature of our study allowed us to build *predictive* models of CDI recurrence, which are not possible with cross-sectional designs. Moreover, by collecting broad gut metabolomic data, we were able to establish that this data can predict CDI recurrence more accurately than microbial composition data. The limited predictive capability of microbial sequencing data could be due to several factors, including lack of data about the status of host processes, poor sensitivity for detecting important low abundance organisms, and the inability to find common signal from diverse bacterial species that perform similar functional roles in the gut. However, it is also possible that predictive computational models specifically tailored to combining microbial compositional and metabolomic data could yield additional information and improve predictive accuracy.

Our findings have implications for design of diagnostic tests and therapeutic interventions for recurrent CDI. We did not find clear differences between recurrers and non-recurrers at the time of CDI diagnosis. Rather, we found that the rate of recovery from dysbiosis was substantially slower in recurrers, with incomplete recovery still evident at two weeks post-CDI antibiotic treatment. Further, we found that at one-week post-CDI antibiotic treatment, increased levels of specific metabolite biomarkers associated with host inflammation accurately predicted future recurrence. Taken together, these findings suggest that diagnostic tests targeting specific metabolites in the first one to two weeks post-CDI treatment may be most accurate and clinically useful. Moreover, by identifying a small set of metabolites that accurately predict recurrence, we have laid the groundwork for developing a feasible clinical test based on a limited biomarker panel that could be cost-effectively measured through targeted LC/MS or other platforms that already exist in clinical laboratories. Our study also uncovered complex and dynamic differences in gut metabolomes, both across time and between recurrers and non-recurrers, which could suggest new avenues for preventing or treating recurrent CDI. For example, we found increased levels of sphingomyelins, sphingolipids, and phospholipids in recurrers prior to the onset of symptoms. These lipids have been found in the guts of late-stage acute CDI in mice, as well as children with IBD and CDI+IBD [10], [17], and may indicate early markers of gut inflammation as *C. difficile* begins to exert pathogenic effects that do not yet cause frank diarrhea or other symptoms. Interestingly, these lipids have recently been shown to be synthesized by common gut bacteria and affect vascular endothelium function and inflammatory responses [42], [43]. Thus, it is possible that rises in these metabolites seen in recurrers at least partially reflect metabolic activity of the microbiome, which could exacerbate development of CDI through modulation of host inflammatory and immune processes. Reduced endocannabinoids in recurrers could similarly involve an interplay between the host and microbiome, as recent evidence suggests that gut microbes regulate endocannabinoids in order to control energy metabolism and gut functions in the host [37].

## Conclusions

We found in our prospective, longitudinal, multi-omic study of CDI recurrence that the microbiomes and metabolomes of participants, while similar immediately before and after initial treatment, diverged rapidly as non-recurrers recovered normal microbiota and metabolic functions and recurrers remained dysbiotic. Our analyses uncovered specific metabolic derangements in participants who experienced subsequent recurrence, including evidence of loss of normal metabolic activities of the gut microbiome, host gut inflammation and cell death, and decreases in anti-inflammatory and immune-modulating compounds. Moreover, we found that differences in specific metabolites in the first two weeks post-CDI antibiotic treatment accurately predicted future recurrence, while microbiome sequencing data did not yield high predictive accuracy. These results suggest that metabolomics may be the more robust modality for evaluating recovery of microbial function. By providing specific candidate predictive biomarkers and expanding our knowledge of the complex metabolic changes preceding recurrence, our findings have implications for development of diagnostic tests and treatments for CDI recurrence

## Methods

### Study design

This prospective, longitudinal cohort study was conducted to study microbiome and metabolome predictors of CDI recurrence. Participants with primary, uncomplicated CDI were identified by positive test results from the Brigham and Women’s Hospital (BWH) Clinical Microbiology Laboratory and recruited from BWH’s inpatient service, as well as two surrounding community hospitals. Participants who were being treated for primary CDI, diagnosed with diarrhea symptoms, and a positive *C. difficile* test by either glutamate dehydrogenase (GDH) and enzyme immunoassay (EIA) toxin or polymerase chain reaction (PCR), were eligible for inclusion. Primary CDI was defined as no episodes of CDI within the past 6 months. Exclusionary criteria included inflammatory bowel disease, inherited or acquired immunodeficiencies, severe or fulminant CDI, or ongoing non-CDI antibiotic use that continued past the CDI antibiotic course. 53 participants were followed from initial diagnosis until recurrence, or for 8 weeks post-treatment if they did not recur. Samples at diagnosis (before initiation of antibiotic treatment for CDI) were available for only 26 participants because of the difficulty in obtaining viable fecal samples from the clinical laboratory workflow. Samples were taken at diagnosis (if available), weekly or bi-weekly for 2 weeks after diagnosis, and then weekly for another 6 weeks, or until recurrence. Recurrence was defined as diarrhea (Bristol stool scale 6 or 7), at least 3 bowel movements daily for 3 days, and a positive GDH and EIA test, in keeping with current guidelines. Because all participants recurred before week 4, samples after week 4 were not analyzed.

### Participant demographics and clinical data

Demographic variables were collected for the 53 participants (Table 1). Weight and height were collected individually and used to calculate participants’ BMI. Significance testing for demographic variables was conducted using Fisher’s exact test for binary variables (sex), the chi-squared test for categorical variables (race, smoking status), and the Wilcoxon-rank-sum test for continuous variables (age, BMI).

### 16S rRNA gene amplicon sequencing

For DNA extraction, all fecal samples were processed with the Zymo Research ZymoBIOMICS DNA 96-well kit according to manufacturer instructions with the addition of bead beating for 20 minutes. The extracted DNA was used for 16S rRNA gene Amplicon sequencing and 16S rRNA qPCR for total bacterial concentration estimation. Amplicon sequencing of the v4 region of the 16S rRNA gene was performed using the previously described protocol in [44] using 515F and 806R primers for PCR along with:

5’-[Illumina adaptor]-[unique bar code]-[sequencing primer pad]-[linker]-[primer] Read 1 (forward primer): AATGATACGGCGACCACCGAGATCTACAC-NNNNNNNN-TATGGTAATT-GT-GTGCCAGCMGCCGCGGTAA

Read 2 (reverse primer): CAAGCAGAAGACGGCATACGAGAT-NNNNNNNN-AGTCAGTCAG-CC-GGACTACHVGGGTWTCTAAT

### LC-MS untargeted metabolomics

LC-MS untargeted metabolomics was performed by Metabolon (Durham, NC USA). After delivery to Metabolon, samples were homogenized in methanol to extract metabolites and then centrifuged to separate the supernatant from debris and precipitates. The supernatant was divided into five aliquots for four analyses plus one extra and then dried using a TurboVap (Zymark). Dried samples were stored overnight under nitrogen gas. Samples were reconstituted and measured with Waters ACQUITY ultra-performance liquid chromatography (UPLC) and Thermo Scientific Q-Exactive high resolution/accurate mass spectrometry (MS), heated electrospray ionization source (HESI-II) and Orbitrap pass analyzer (35,000 mass resolution). Samples were analyzed in the following four different ways: (1) elution w/ C18 column (Waters UPLC BEH C18-2.1×100mm, 1.7um) in positive-ion mode with water/methanol gradient mobile phase containing 0.05% perfluorpentanoic acid (PFPA) and 0.1% formic acid (FA), (2) as in (1), except w/ water/acetonitrile/methanol gradient mobile phase containing 0.05% PFPA and 0.01% FA, (3) elution w/ a separate C18 column in negative-ion mode w/ water/methanol gradient mobile phase containing 6.5 mM ammonium bicarbonate, pH 8, and (4) elution w/ HILIC column (Waters UPLC BEH amide 2.1×150mm, 1.7um) in negative-ion mode w/ water/acetonitrile gradient mobile phase containing 10 mM ammonium formate, pH 10.8. The MS analysis was performed as dynamic exclusion, altering between MS and data-dependent MS^n^ scans with a scan range of 70-1000 m/z. Data extraction, peak identification, quality control, and annotation were performed by Metabolon’s proprietary software.

### Short chain fatty acid profiling

Volatile SCFAs were quantified as described in [45]. Acidified internal standards with 100uL of either ethyl anhydrous or boron trifluoride-methanol was added to 100uL of supernatant from homogenized cecal contents. Chromatographic analyses were carried out on an Agilent 7890B system with a flame ionization detector (FID). Chromatogram and data integration were done using the OpenLab ChemStation software (Agilent Technologies, Santa Clara, CA). SCFAs were identified by comparing their specific retention times relative to the retention time in the standard mix. Concentrations were determined as mM of each SCFA per gram of sample for the raw cecal/fecal material. The Agilent platform cannot discriminate between isovalerate and 2-methylbutyrate, and so these are reported as a single peak value.

### 16S rRNA gene amplicon data analysis

#### Bioinformatics

Sequencing generated 4,605,740 total reads and 97,994 average reads per sample. The paired-end Fastq files were truncated, filtered, denoised, and merged using the dada2 pipeline and filtering parameters identical to the dada2 tutorial [46]. Our analysis found 2509 unique amplicon sequencing variants (ASVs) in the dataset, and taxonomy was assigned using the dada2 RDP and Silva reference databases [47], [48]. If taxa assignments between the two databases disagreed, the taxa assignment from the RDP database was used.

#### Alpha and beta diversity

Prior to calculating alpha and beta diversity, relative ASV abundance was calculated by dividing each ASV’s counts by the total number of counts in a sample. Using ASV relative abundance, we calculated the alpha diversity (Chao1) at pre-treatment, week one, week two, week three, and week four using scikit-bio (skbio.diversity.alpha.chao1) [24]. Significant differences in intra-group alpha diversity over time and inter-group alpha diversity at pre-treatment, week one, and week two were tested using the Mann-Whitney U test. A one-sided test was used to test the hypothesis that alpha diversity of both groups decreased during antibiotic treatment and then recovered, and the hypothesis that non-recurrent participants would have higher alpha diversity. Beta diversity was calculated at pre-treatment, week one, week two, week three, and week four from the Bray-Curtis dissimilarities (calculated using scipy.spatial.distance.pdist) of relative ASV abundances between each subject. To visualize the dissimilarity of outcome groups at each timepoint and the intra-group dissimilarity between timepoints, we performed multi-dimensional scaling (using scikit-learn.manifold.MDS) on the Bray-Curtis dissimilarities. We used permutational multivariate analysis of variance (PERMANOVA) (skbio.stats.distance.permanova) with 999 permutations to assess the significance of inter and intra group dissimilarities at pre-treatment, week one, and week two.

#### Filtering

Prior to differential abundance analysis, ASVs were filtered to remove rare taxa. We included ASVs present with >10 counts and in ≥10% of participants in either pre-treatment, week one, or week two. This resulted in 237 ASVs post-filtering.

#### Differential abundance analysis

After filtering, differential abundance analyses between recurrers and non-recurrers at pre-treatment, week one, and week two were performed using the DESeq function within the DESeq2 package [49]. Because every ASV in the dataset contained zeros, we pre-computed the geometric means and then the size factors using the estimateSizeFactors function within DESeq2. Intra-group differential abundance analysis was also performed between pre-treatment and week one, and between week one and week two, for both recurrers and non-recurrers using the same procedure in DESeq2. All differential abundance analyses were followed by the Benjamini-Hochberg correction for multiple hypotheses [50]. The relative abundances of ASVs that were significantly different between recurrers and non-recurrers at pre-treatment, week one, or week two are shown in Figure 2 on a logarithmic scale, along with the phylogenetic relationships of these ASVs (found with methods detailed below). In this figure, recurrers and non-recurrers at each timepoint are clustered hierarchically using scipy.cluster.hierarchy with optimal ordering and ‘average’ distance.

#### Phylogenetic placement

To further clarify phylogenetic relationships between ASVs of interest, we built a reference tree and then performed phylogenetic placement of ASVs. For the reference tree, all typed, isolated strains of good quality that were longer than 1200 base pairs were downloaded from the RDP bacteria and archaea datasets [47]. Reference sequences were then aligned using the RDP aligner. The reference sequences were then filtered to remove: (1) sequences with unaligned lengths ≥1600 bp and, (2) sequences with rare insertions (defined as a base pair in a position where there were 5 or less sequences with un-gapped base pairs in that position). Filtered reference sequences were then realigned using the same RDP aligner. A reference tree was constructed using FastTree version 2.1.7 SSE3 with the general-time-reversible maximum likelihood option [51]. Pplacer v1.1.alpha19 with default settings [52] was then used to place query ASVs onto the reference tree.

#### Enrichment analysis

Enrichment analyses were performed on the ASVs found in each differential abundance analysis with FDRs<0.05 (Table S3). For a given family A, we tested if the family was significantly overrepresented in differentially abundant ASVs using the hypergeometric probability distribution:

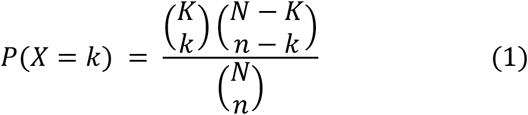

Here, *N* is the total number of (post-filtering) ASVs, *K* is the subset of *N* in family A, *n* is the number of differentially abundant ASVs, and *k* is the subset of *n* in family A. To prevent false positives due to small family sizes, we did not test (1) families that had too few ASVs in the total post-filtering set (K≤3) or (2) families that had too few ASVs in the differentially abundant subset (k≤2). For all families large enough to pass the filter, *p*-values were computed using the hypergeometric test, and the Benjamini-Hochberg procedure was using to correct for multiple hypothesis testing [50].

### LC-MS untargeted metabolomics data analysis

“OrigScale” data returned by Metabolon was used in all analyses described in this manuscript; these data represent values normalized in terms of raw area counts.

#### Ordination analyses

To assess inter-group dissimilarity at each timepoint and intra-group dissimilarity between timepoints, we computed matrices using Spearman rank correlation on the unfiltered and untransformed metabolomic data. We used PERMANOVA (skbio.stats.distance.permanova) with 999 permutations to test the significance of differences (Table S1).

#### Filtering

LC-MS untargeted metabolomics measured 1387 metabolites. To retain only metabolites with sufficient prevalence and variance across time or participants, we included metabolites with: (1) non-zero values in ≥25% of participants in either pre-treatment, week one, or week two samples, and (2) co-efficient of variations in the top 50^th^ percentile in either pre-treatment, week one, or week two samples. These criteria resulted in 760 metabolites post-filtering.

#### Univariate analysis

Before univariate analyses, metabolite values were log transformed (after adding a small positive number, 10% of the minimum non-zero value in the dataset) to all values, and standardized to have a mean of zero and a standard deviation of one. After filtering and transforming the metabolic data, we performed statistical testing using Student’s *t*-test followed by Benjamini-Hochberg correction for multiple hypotheses testing [50].

#### Enrichment analysis

Enrichment analyses were performed on the metabolites found in univariate analysis with FDRs<0.05, with an analogous method as used for enrichment analysis of the ASVs. We used the hypergeometric test with Benjamini-Hochberg [50] multiple hypothesis correction to assess whether pre-specified groups (super pathways and sub-pathways) were significantly over-represented in the differentially abundant metabolites. As with the ASV enrichment analysis, we did not perform hypothesis testing on super and sub-pathways with: (1) too few metabolites in the total post-filtering set (K≤3) or, (2) too few metabolites in the differentially abundant subset (k≤2).

### Short chain fatty acid data analysis

SCFA profiling found eight SCFAs in the analyzed samples: acetate, propionate, isobutyrate, butyrate, isovalerate/2-methylbutyrate, valerate, heptanoate, isocaproate, and caproate. Heptanoate was removed from subsequent analyses due to only being present in one sample. Caproate was present in only two samples and was also removed from further analyses. The remaining seven SCFAs were log transformed and standardized analogously to the untargeted metabolomics data prior to univariate analysis; univariate analyses were also performed analogously to those on the untargeted metabolomics data.

### Predictive modeling

The following data sources were used as input to predictive modeling methods: (1) clinical variables found to be associated with recurrence in prior studies (age, previous PPI use, antibiotic treatment regiment, and CDI diagnostic test), (2) 16S rRNA amplicon sequencing (ASVs) from pre-treatment, week 1, or week 2 samples, (3) untargeted metabolomics data from pre-treatment, week 1, or week 2 samples, (4) SCFAs profiles from pre-treatment, week 1, or week 2 samples, and (5) data sources 1-4 combined. In each predictive model, training datasets were filtered with the same criteria described for univariate analyses. Metabolites and SCFAs were log-transformed and standardized, and ASV relative abundances were transformed with the centered log ratio and then standardized. Continuous clinical variables (i.e., age) were log-transformed and standardized.

Relevant predictive features were identified through a nested leave-one-out cross-validation procedure (described in detail below for each method). To summarize the results for each feature, we report the median and 95% interval over the folds (i.e., regression coefficients for logistic and cox regression, feature importances for random forests). We deem features significant if the 95% cross-validated odds-ratio/feature-importance intervals did not contain 1.0, and marginally-significant if the 75% cross-validated interval did not contain 1.0. The code to reproduce these analyses can be found in https://github.com/gerberlab/cdiff_paper_analyses.

#### Logistic regression

Logistic regression models were fit using scikit-learn’s (v0.24.2) logistic regression function with L1 lasso regularization, balanced classes, and a liblinear solver. We used nested leave-one-out cross validation to find the optimal L1 hyperparameter, performing a grid search over a range of 200 values from the maximum lambda value (i.e., the value that resulted in all zero coefficients) to 0.1% of the maximum lambda value. Performance in the inner loop was evaluated by area under the receiver operator curve (AUC) score calculated from the predictions of all the held-out samples. To reduce overfitting, the inner loop performances were smoothed using a n=5 moving average, and the optimal L1 hyperparameter was that which resulted in the highest value on the smoothed performance curve. After choosing the best L1 hyperparameter, the model’s predictive capability was evaluated by its leave-one-out cross validated AUC score. Variance estimates of model performance and regression coefficients were calculated from the cross-validation folds.

#### Random forest

Random forest models were fit using scikit-learn’s (v0.24.2) random forest classifier. We performed a nested leave-one-out cross validation procedure with grid search, to determine the number of estimators (50 or 100), the maximum features to subsample at each split (the total number of features or the square root of the number of features), the minimum samples required to split an internal node (2 or 9) and the minimum samples required to split a leaf node (1 or 5). All other parameters were set to their default values except for class weight (‘balanced’) and out of box score (True). The feature importances were calculated with the impurity-based feature importance, or the Gini importance, using the feature_importance attribute of the fitted model.

Model performance and feature importance statistics were calculated from the cross-validation folds.

#### Cox regression

Cox regression models were fit using scikit-survival’s (v0.15.0) Coxnet Survival Analysis function with L1 regularization. We used a similar nested cross validation as described for our logistic regression analyses to optimize the L1 parameter, searching over a range of 200 values from the maximum lambda value (i.e., the value that resulted in all zero coefficients) to 0.01% of the maximum lambda value. We evaluated both the inner and outer loops of the survival analysis using the concordance index (CI). Rather than leave-one-out cross validation, we used a leave-two-out method, where all left out pairs had at least one recurrer, to calculate the CI. In this formulation (mathematically equivalent to the standard definition of CI), CI is computed by dividing the number of times a pair was ordered correctly by the number of times a pair ordering was attempted. Variance estimates of model performance were calculated from the cross-validation folds.

## Data Availability

The sequencing datasets generated and/or analyzed during the current study are available in the SRA repository, accession number PRJNA772946
Data for reproducing all figures and analyses are available in Zenodo.
All other datasets supporting the conclusions of this article are included within the article and its additional files.
Code to reproduce the analysis and figures in this paper is publicly available in the Github

https://dataview.ncbi.nlm.nih.gov/object/PRJNA772946

https://doi.org/10.5281/zenodo.5703428

https://doi.org/10.5281/zenodo.5718583

https://github.com/gerberlab/cdiff_paper_analyses

## Declarations

All study participants signed informed consent prior to study procedures. The protocol was approved by the institutional review board at Brigham and Women’s Hospital (#2014P000272).

### Consent for publication

Not applicable

### Availability of data and material

The sequencing datasets generated and/or analyzed during the current study are available in the SRA repository, accession number PRJNA772946, https://dataview.ncbi.nlm.nih.gov/object/PRJNA772946. Data for reproducing all figures and analyses are available in Zenodo at https://doi.org/10.5281/zenodo.5703428 and https://doi.org/10.5281/zenodo.5718583.

All other datasets supporting the conclusions of this article are included within the article and its additional files.

Code to reproduce the analysis and figures in this paper is publicly available in the Github project cdiff_paper_analyses at at https://github.com/gerberlab/cdiff_paper_analyses. Code is platform independent and written in python (3.7.10) and R (4.1.0). All R and python packages and version numbers can be found in the Github repo readme.md file.

### Funding

JA: Junior Faculty Development Award from the American College of Gastroenterology. LB: BWH Precision Medicine Institute, Hatch Family Foundation, P30 DK056338.

TEG: NIH NIGMS R35GM143056, NIH NIAID R21AI154075.

GKG: NIH NIGMS R01 GM130777, Harvard Catalyst, BWH Precision Medicine Institute, BWH President’s Scholar Award.

### Authors’ contributions

JJD analyzed and interpreted the data, wrote the software to analyze the data, and wrote the manuscript. JA designed and oversaw the clinical study, interpreted the data, and edited the manuscript. TEG analyzed and interpreted the data, designed analyses methods, and edited the manuscript. EM coordinated the clinical study. MD prepared and analyzed microbiological samples. LB designed the clinical study, oversaw experimental data generation, interpreted the data, and edited the manuscript. GKG designed the clinical study, analyzed and interpreted the data, and wrote the manuscript. All authors read and approved the final manuscript.

## Acknowledgements

David Kaplan provided advice on phylogenetic tree building and placement. Figure 1 was created with BioRender.com.

## Supplemental Figures

**Supplemental Figure S1:**
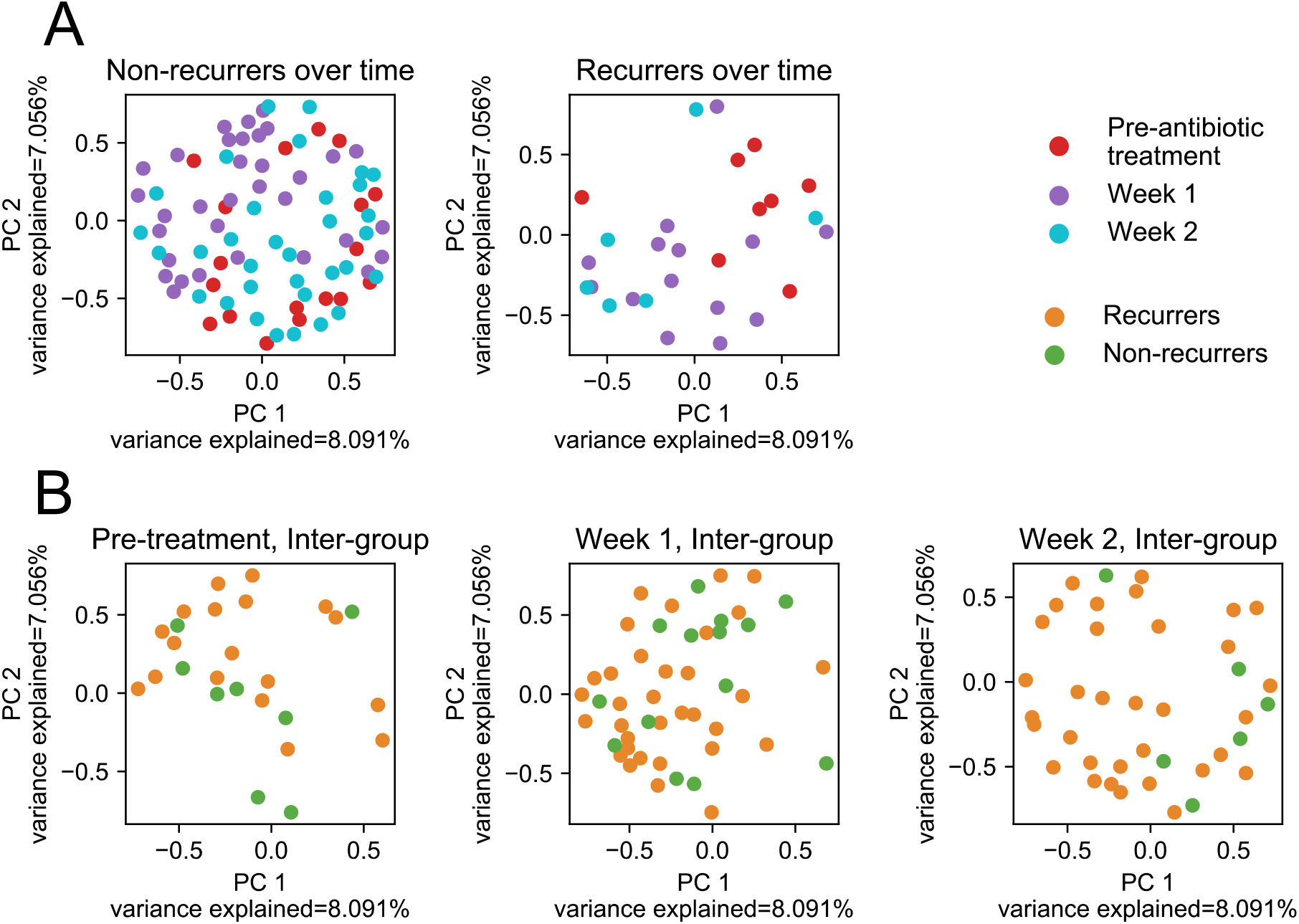
Gut microbiome community structure significantly changed over time within recurrers or non-recurrers and was significantly different between the groups at week two. Beta diversity with the Bray-Curtis dissimilarity measure was used to assess overall microbiome community structure; Principal Coordinate Analysis (PCoA) was used to visualize results. **(A)** Beta diversity changed significantly over time within groups. Differences were significant for non-recurrers from pre-treatment to week one (p=10^−3^) and from week one to week two (*p*=10^−3^). For recurrers, differences were significant from pre-treatment to week one (*p*=3×10^−3^). **(B)** Beta diversity was significantly different between recurrers and non-recurrers at week two (p=10^−2^); differences at other time-points were not significant.

**Supplemental Figure S2:**
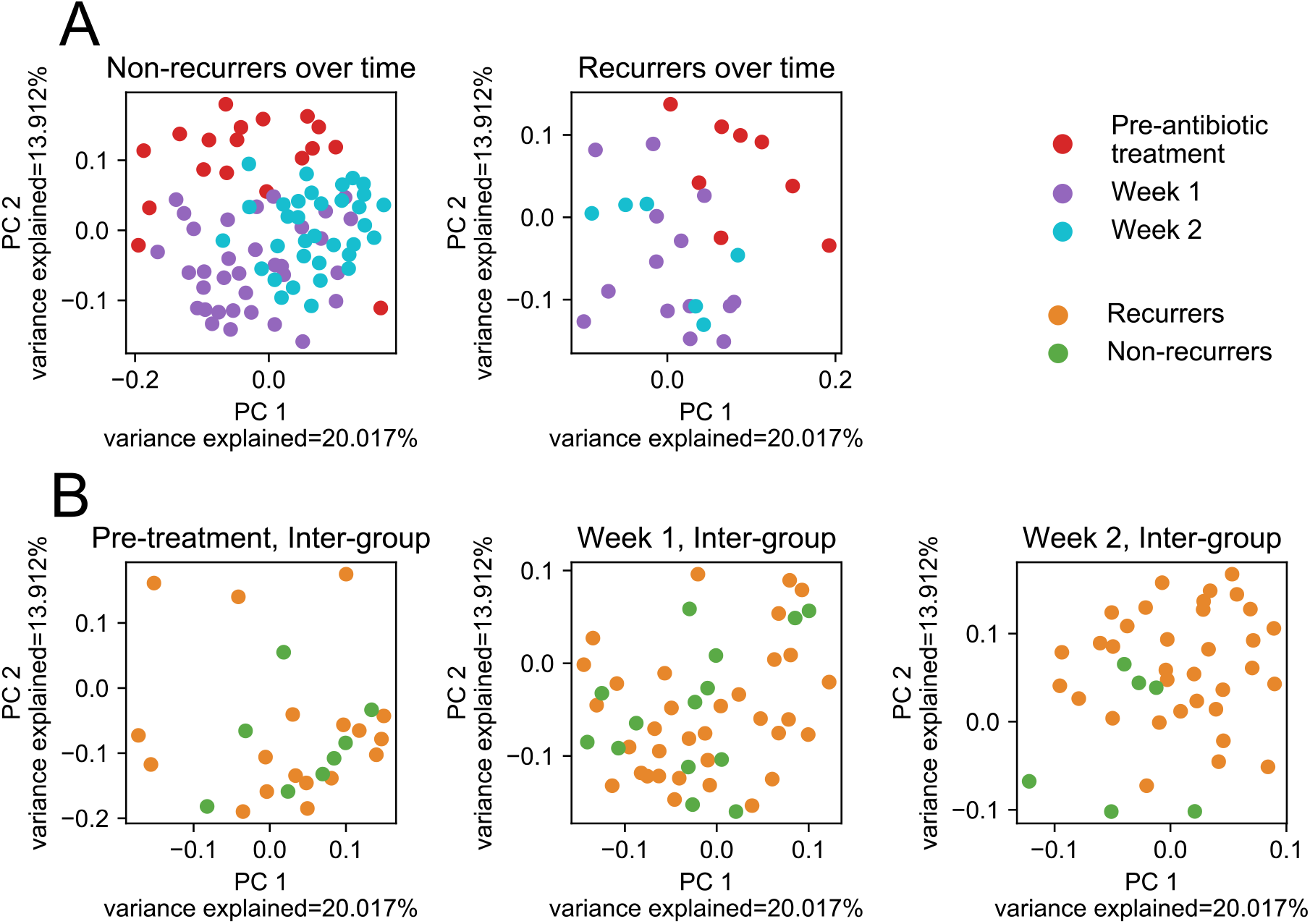
Gut metabolome structure significantly changed over time within recurrers or non-recurrers and was significantly different between the groups at week two. Ordination analysis using Spearman rank correlation was used to assess overall metabolome structure; Principal Coordinate Analysis (PCoA) was used to visualize results. **(A)** Metabolome structure changed significantly over time within groups. Differences were significant for non-recurrers from pre-treatment to week one (p=10^−3^) and from week one to week two (*p*=10^−3^). For recurrers, differences were significant from pre-treatment to week one (*p*=10^−3^). **(B)** Metabolome structure was significantly different between recurrers and non-recurrers at week two (p=10^−3^); differences at other time-points were not significant.

**Supplemental Figure S3:**
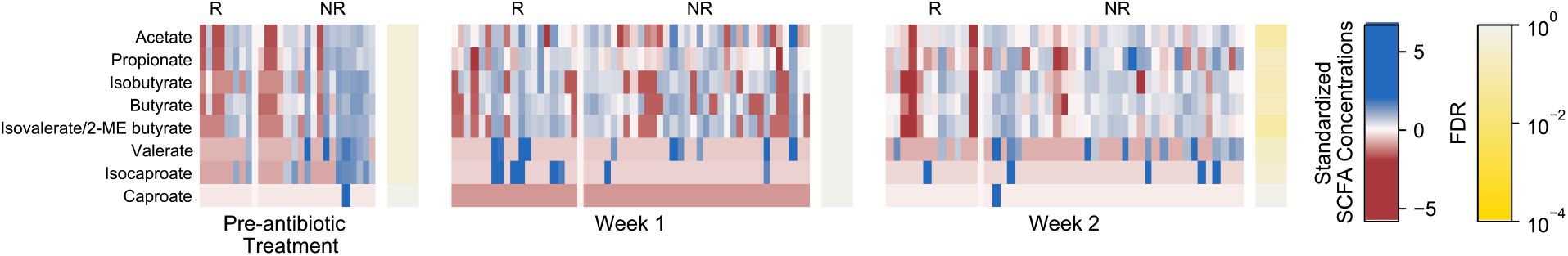
A border-line significant trend was observed of higher levels of fecal acetate and isovalerate short-chain fatty acids at week two in participants who did not recur with *C. difficile*. Log-transformed and standardized concentrations of the short-chain fatty acids measured in fecal samples are shown. Levels of acetate (FDR=0.07) and isovalerate/2-ME butyrate (FDR=0.07) were higher in non-recurrent (NR) versus recurrent (R) participants.

